# Prevalence and population attributable fractions of potentially modifiable risk factors for dementia in Canada: a cross-sectional analysis of the Canadian Longitudinal Study on Aging

**DOI:** 10.1101/2024.04.06.24305404

**Authors:** Yasaman Dolatshahi, Alexandra Mayhew, Megan E. O’Connell, Teresa Liu-Ambrose, Vanessa Taler, Eric E. Smith, David B. Hogan, Susan Kirkland, Andrew P. Costa, Christina Wolfson, Parminder Raina, Lauren Griffith, Aaron Jones

## Abstract

**Background:** Identification and assessment of modifiable risk factors for dementia is a public health priority in Canada and worldwide. We investigated the prevalence and population attributable fraction (PAF) of 12 potentially modifiable risk factors for all-cause dementia in middle-aged and older Canadians.

**Methods:** We conducted a cross-sectional study of data from the Comprehensive cohort of the Canadian Longitudinal Study on Aging, a national sample of 30,097 individuals between the ages of 45 and 85 at baseline (2011-2015). Risk factors and associated relative risks were taken from a highly cited systematic review published by an international commission on dementia prevention. We estimated the prevalence of each risk factor using sampling weights to be more generalizable to the Canadian population. Individual PAFs were calculated both crudely and weighted for communality, and combined PAFs were calculated with methods reflecting both multiplicative and additive interaction assumptions. Analyses were additionally performed stratified by household income and were repeated at CSLA’s first three-year follow-up (2015-2018).

**Results:** The most prevalent risk factors at baseline were physical inactivity (63.8%; 95% CI, 62.8% – 64.9%), hypertension (32.8%; 31.7% – 33.8%), and obesity (30.8%; 29.7% – 31.8%). The highest crude PAFs were for physical inactivity (19.9%), traumatic brain injury (16.7%), and hypertension (16.6%). The highest weighted PAFs were for physical inactivity (11.6%), depression (7.7%), and hypertension (6.0%). We estimated that the 12 risk factors combined accounted for 43.4% (37.3%-49.0%) of dementia cases assuming weighted multiplicative interactions and 60.9% (55.7%-65.5%) assuming additive interactions. There was a clear gradient of increasing prevalence and PAF with decreasing income for 9 of the 12 risk factors.

**Interpretation:** The findings of this study can inform individual and population-level dementia prevention strategies in Canada, focusing efforts on risk factors with the largest impact on the number of dementia cases. Differences in the impact of individual risk factors between this study and other international and regional studies highlight the importance of tailoring national dementia strategies to the local distribution of risk factors.

## Introduction

Dementia is a clinical syndrome marked by typically progressive cognitive decline, affecting memory, perceptual-motor skills, language, mood, behaviour, and the ability to perform activities of daily living ^1–3^. In Canada, the number of people living with dementia was estimated to be 597,000 in 2020 and is expected to increase to 1.7 million by 2050^4,5^. The increase in the number of individuals living with dementia will also greatly affect families and care partners who provide extensive hours of unpaid care^5^. Total health system and out-of-pocket costs for dementia care in Canada are currently estimated to be $10 billion and the rising number of individuals living with dementia will have a substantial effect on health and social care systems^4,5^. Effective treatment options for dementia are limited, placing an emphasis on the importance of prevention^1,5^. The effectiveness of prevention strategies, however, is dependent on identifying and understanding potentially modifiable risk factors associated for dementia as well as their prevalence and distribution within a population^5^.

The 2020 report of the Lancet Commission on dementia prevention, intervention, and care identified 12 potentially modifiable risk factors^6^: less education, hearing loss, traumatic brain injury (TBI), hypertension, excessive alcohol consumption, obesity, smoking, depression, social isolation, physical inactivity, air pollution, and diabetes. They estimated that each risk factor accounted individually for between 2.0% to 22.0% of dementia cases and together accounted for 40% of dementia cases worldwide^6^. The distribution of modifiable risk factors for dementia has been has been examined in detail in other countries to inform national and regional public health strategy^6–8^. However, research using national Canadian data has been lacking despite the importance of research on modifiable risk factors being highlighted in Canada’s national dementia strategy^9^. In addition, while lower socio-economic status is associated with increased dementia incidence, little work has examined how the prevalence of modifiable risk factors for dementia varies by income^10^.

The objective of this study is to examine the prevalence and impact of potentially modifiable risk factors for dementia in Canada based on data from the Canadian Longitudinal Study on Aging (CLSA). Our specific aims are to estimate 1.) the prevalence of 12 potentially modifiable risk factors for all-cause dementia in the Canadian middle-aged and older adult population and 2.) the proportion of dementia cases attributable to each risk factor, i.e. the population attributable fraction (PAF), alone and in combination. Secondarily, we will examine how the distribution of these risk factors varies by household income.

## Methods

### Setting and study type

We conducted a cross-sectional study of middle-aged and older adults in Canada.

### Data Sources

The Canadian Longitudinal Study on Aging (CLSA) is a national, stratified sample of 51,338 Canadian residents between the ages of 45 and 85 at baseline recruitment (2011-2015). Participants in CLSA were recruited via three sampling frames (the Canadian Community Health Survey, random-digit dialing, and provincial health registries). The CLSA is comprised of two cohorts, Tracking and Comprehensive. This study uses data only from the Comprehensive cohort as the calculation of all 12 risk factors of interest is not possible on the Tracking cohort. Comprehensive cohort participants (n=30,097 at baseline) were randomly selected within proximity of 11 data collection sites in 7 provinces (Newfoundland and Labrador, Nova Scotia, Ontario, Quebec, Manitoba, Alberta, British Columbia) with data collected in-person during home interviews and at the data collection sites. The methodology and sampling strategy of the CLSA has been fully described elsewhere^11,12^. Air quality data were collected and provided by the Canadian Urban Environmental Health Research Consortium and Health Canada and linked to participants by postal code and interview date^13^.

### Study participants

We included all Comprehensive cohort participants at baseline (2011-2015) and follow-up 1 (2015-2018). At the time of recruitment CLSA participants had to be able to complete interviews in English or French and be able to participate in the study on their own. Residents of the three Canadian territories, First Nations reserves, or institutions; full-time members of the Canadian Armed Forces; and individuals unable to consent due to cognitive impairment were excluded from baseline recruitment.

### Participant characteristics

To characterize the study sample, we extracted self-reported data on demographics, income, number of chronic conditions, and general health. Income measures were based on total household income and categorized as implemented in the CLSA: less than $20,000, $20,000 - $49,999, $50,000-$99,999, $100,000 - $149,999 and $150,000 or higher^14^.

### Modifiable risk factors for dementia

The definitions for the 12 modifiable risk factors associated with all-cause dementia were guided where possible by the 2020 Lancet Commission Report, other published work on modifiable risk factors for dementia, and the 2020 Canadian federal air quality guidelines^6,7,15,16^. The specific definitions for the risk factors are listed in Table 1. A comparison of these definitions with similar studies can be found in Supplemental Table S1. All definitions except for air quality, hearing loss, and obesity were defined using CLSA self-report interview data. Air quality was based on national land-use regression models and satellite-derived measures^17–21^. Hearing loss was calculated using pure tone audiometry. Obesity was calculated using in-person BMI measurements. The relative risks associated with each risk factor were taken from the meta-analyses reported by the Lancet Commission^6,22^.

**Table 1.** Definitions of potentially modifiable risk factors for dementia.

| Risk Factor | Definition |
| --- | --- |
| Less Education | Less than secondary school graduation |
| Hearing Loss | Pure-tone audiometry: Average loss in better ear > 25 dB measured at 0.5, 1, 2, and 4 kHz |
| Traumatic Brain Injury | Lifetime history of head injury resulting in loss of consciousness, loss of memory, or dazedness |
| Hypertension | Self-reported medical diagnosis of high blood pressure or hypertension |
| Excessive alcohol consumption | Self-reported frequency of drinking more than 15 alcoholic drinks for males and 10 for females per week in the past 12 months |
| Obesity | Measured body mass index (BMI) $\geq 30$ kg/m<sup>2</sup> |
| Smoking | Self-reported current smoking (at least one cigarette every day for the past 30 days) |
| Depression | CESD-10 score $\geq 10$ |
| Physical inactivity | Self-reported physical activity of less than 75 minutes of vigorous activity or less than 150 minutes of combined moderate and vigorous activity per week |
| Diabetes | Self-reported medical diagnosis of diabetes, borderline diabetes, or high blood sugar |
| Social isolation | Self-reported frequency of participation in family/friend activities outside of household, religious activities, physical activities with other, educational/cultural activities, association activities, volunteer/charity work, and other recreational activities all less than once per week |
| Air pollution | Nitrogen dioxide (NO<sub>2</sub>): Annual average $\geq 17$ ppb *or*<br>Fine particle matter (PM<sub>2.5</sub>): 3-year annual average $\geq 8.8$ µg/m<sup>3</sup> *or*<br>Sulfur dioxide (SO<sub>2</sub>): 3-year annual average $\geq 5$ ppb |

### Statistical Analysis

We constructed a descriptive profile of CLSA Comprehensive cohort participants at baseline, summarizing demographics, socioeconomic factors, and health status variables by measures of frequency, central tendency, and dispersion. In the primary analysis, the prevalence and 95% confidence interval of each modifiable risk factor was calculated with the use of sampling weights, which were designed to generalize to the target population of Canadian middle-aged and older adults. CLSA sampling weights were calibrated using age, sex, and education (within geographical regions)^23^. Within each calculation, individuals with missing data were removed on a per factor basis. For 10 of the risk factors, the proportion of missing values was minimal, ranging from 0% to 0.63%. Missingness was somewhat higher for physical inactivity (4.4%) and hearing loss (5.0%).

The crude PAF for each risk factor was estimated by Levin’s formula^24^. Additionally, following the methodology introduced by Norton et al., we produced weighted PAFs to account for overlap between risk factors^25^. We generated communality weights by conducting principal component analysis on the tetrachoric correlation matrix of the 12 risk factors, extracting only components with eigenvalues > 1. The communality for each risk factor, which represents the degree to which it overlaps the other risk factors, was calculated as the sum of the squared factor loadings. The corresponding weight for each risk factor, representing the factor’s uniqueness, was the complement of the communality.

We calculated two combined PAFs with different assumptions regarding the form of interactions between risk factors. The first is a widely-used method developed by Norton et al. that multiplicatively combines the individual communality-weighted PAFs^6,7,25^. This method was developed to yield more plausible estimates of PAFs involving multiple risk factors than other methods that multiplicatively combined crude PAFs^25,26^. The second method of calculating a combined PAF assumes additive interactions between risk factors and has been suggested as an alternative to the communality-weighted multiplicative approach^26^. A technical appendix with all formulae can be found in Supplemental Table S2. We derived confidence intervals for all PAFs using a two-step bootstrapping approach to account for uncertainty in both the risk factor prevalence and relative risk estimate^26^. In this process we first generated 10,000 replicate samples with replacement and then within each replicate we parametrically sampled relative risks for each risk factor based on their reported confidence intervals.

We repeated all analyses within household income categories. We reported the prevalence and combined PAFs within each category of income and plotted the prevalence by income to facilitate comparison. Data were analyzed using the R programming language with the packages “survey” and “psych”^27,28^.

### Sensitivity Analysis

We repeated the main analysis at the first follow-up data collection point in CLSA, which was approximately three years after baseline (2015-2018). To assess sensitivity to the use of sampling weights, we repeated all analysis using the crude prevalence for each risk factor. Finally, we also examined prevalence and PAF of each risk factor at baseline by sex.

### Ethics

This study received ethics approval from the Hamilton Integrated Research Ethics Board (15408).

## Results

The baseline CLSA Comprehensive cohort included 30,097 participants and was 50.9% female (Table 2) The median age was 62 years with first and third quartiles of 54 and 71 years. A small proportion of participants (8.1%) resided in rural areas. The participants were primarily white only (94.4%), followed by multiple identities (1.5%), South Asian only (0.9%) and Black only (0.7%). A small proportion (5.6%) of the sample reported an annual household income below $20,000, 22.6% reported between $20,000 and $49,999, 35.2%, between $50,000 and $99,999, 19.6% between $100,000 and $149,999, and 17.0% reported an annual household income of $150,000 or above. While nearly all participants (91.8%), reported having at least one chronic condition, over half (61.5%) of participants reported their general health to be “Excellent” or “Very Good” and only 1.3% reported that it was “Poor”.

**Table 2:** Characteristics of CLSA Comprehensive cohort at baseline.

| Characteristic | CLSA Comprehensive Cohort<br>(n=30,097) |
| --- | --- |
| **Sociodemographic** |  |
| Sex, Female | 15,320 (50.9%) |
| Age, yrs., median (Q1, Q3) | 62 (54, 71) |
| Age, yrs. |  |
| 45-54 | 7595 (25.2%) |
| 55-64 | 9856 (32.7%) |
| 65-75 | 7362 (24.5%) |
| 75+ | 5284 (17.6%) |
| Rural resident | 2424 (8.1%) |
| Cultural / Racial Background |  |
| White only | 28,372 (94.4%) |
| Black only | 221 (0.7%) |
| Chinese only | 212 (0.7%) |
| South Asian only | 271 (0.9%) |
| Multiple | 447 (1.5%) |
| Other | 544 (1.8%) |
| Total household Income (CAD) |  |
| <\$19,999 | 1566 (5.6%) |
| \$20,000 - \$49,999 | 6360 (22.6%) |
| \$50,000 - \$99,999 | 9907 (35.2%) |
| \$100,000 - \$149,999 | 5524 (19.6%) |
| >\$150,000 | 4799 (17.0%) |
| **Health Status** |  |
| At least one chronic condition | 27,633 (91.8%) |
| Self-rated general health |  |
| Excellent | 5995 (19.9%) |
| Very good | 12,420 (41.3%) |
| Good | 8877 (29.5%) |
| Fair | 2315 (7.7%) |
| Poor | 467 (1.6%) |
| Self-rated healthy aging |  |
| Excellent | 5579 (18.6%) |
| Very good | 12,899 (42.9%) |
| Good | 9023 (30.0%) |
| Fair | 2137 (7.1%) |
| Poor | 398 (1.3%) |

### Prevalence of risk factors

Using sample-weighted proportions, physical inactivity was the most prevalent risk factor (63.8%; 95% CI, 62.8% – 64.9%) followed by hypertension (32.8%; 31.7% – 33.8%), obesity (30.8%; 29.7% – 31.8%) and traumatic brain injury (23.9%; 22.9% – 24.8%) (Table 3). Smoking (9.6%; 8.9% – 10.3%), excessive alcohol consumption (11.4%; 10.8% - 12.0%), and hearing loss (15.7%; 14.9% – 16.5%) were the least common risk factors in the Canadian middle-aged and older adult population.

**Table 3.** The weighted prevalence, relative risk, communality, and population attributable fractions (PAF) of dementia risk factors in the CLSA comprehensive cohort at baseline (2011-2015, n=30,097).

| **Risk Factor** | **Prevalence, %**<br>**(95% CI)** | **Relative risk**<br>**(95% CI)** | **Communality**<br>**%** | **Crude, PAF,**<br>**%** | **Weighted, PAF,**<br>**%** |
| --- | --- | --- | --- | --- | --- |
| Less Education | 17.4 (16.1 – 18.6) | 1.59 (1.26 – 2.01) | 54 | 9.3 (4.3, 15.0) | 4.3 (2.0, 6.9) |
| Hearing Loss | 15.7 (14.9 – 16.5) | 1.94 (1.38 – 2.73) | 74 | 12.9 (5.7, 21.5) | 3.3 (1.5, 5.6) |
| TBI | 23.9 (22.9 – 24.8) | 1.84 (1.54 – 2.20) | 71 | 16.7 (11.4, 22.3) | 4.8 (3.3, 6.5) |
| Hypertension | 32.8 (31.7 – 33.8) | 1.61 (1.16 – 2.24) | 64 | 16.6 (4.9, 28.9) | 6.0 (1.8, 10.4) |
| Excessive Alcohol Consumption | 11.4 (10.8 – 12.0) | 1.18 (1.06 – 1.31) | 82 | 2.0 (0.7, 3.4) | 0.4 (0.1, 0.6) |
| Obesity | 30.8 (29.7 – 31.8) | 1.60 (1.34 – 1.92) | 64 | 15.6 (9.4, 22.1) | 5.6 (3.4, 8.0) |
| Smoking | 9.6 (8.9 – 10.3) | 1.59 (1.15 – 2.20) | 61 | 5.4 (1.5, 10.3) | 2.1 (0.6, 4.0) |
| Depression | 18.5 (17.6 – 19.4) | 1.90 (1.55 – 2.33) | 46 | 14.3 (9.3, 19.8) | 7.7 (5.0, 10.7) |
| Social Isolation | 17.5 (16.6 – 18.5) | 1.57 (1.32 – 1.85) | 48 | 9.1 (5.4, 13.0) | 4.7 (2.8, 6.8) |
| Physical Inactivity | 63.8 (62.8 – 64.9) | 1.39 (1.16 – 1.67) | 42 | 19.9 (9.2, 29.6) | 11.6 (5.4, 17.1) |
| Diabetes | 17.4 (16.5 – 18.2) | 1.54 (1.33 – 1.79) | 59 | 8.6 (5.4, 12.1) | 3.5 (2.2, 4.9) |
| Air Pollution | 19.0 (18.0 – 20.0) | 1.09 (1.07 – 1.11) | 52 | 1.7 (1.3, 2.1) | 0.8 (0.6, 1.0) |
| **Combined Factors** | **Combined PAF, %** |  |  |  |  |
| Weighted Multiplicative | 43.4 (37.3, 49.0) |  |  |  |  |
| Additive | 60.9 (55.7, 65.5) |  |  |  |  |

### Populational attributable fractions

Physical inactivity (19.9%; 9.2%-29.6%), hypertension (16.6%, 4.9%-28.9%) and TBI (16.7%, 11.4%-22.3%) had the highest crude PAFs (Table 3). Conversely, air pollution (1.7%; 1.3% −2.1%), excessive alcohol consumption (2.0%; 0.7-3.4%), and smoking (5.4%; 1.5%-10.3%), had the lowest crude PAFs. In the principal component analysis, we extracted five factors with eigenvalues > 1, representing 60% of the total variation. The communality values for each risk factor varied from 42% to 82%. Accounting for overlap between risk factors, the weighted PAFs indicated that physical inactivity (11.6%; 5.4%-17.1%), depression (7.7%; 5.0%-10.7%), and hypertension (6.0%; 1.8%-10.4%) were associated with the most dementia cases while excessive alcohol consumption (0.4%; 0.1%-0.6%), air pollution (0.8%; 0.6%-1.0%), and smoking (2.1%; 0.6%-4.0%), were responsible for the fewest cases. The combined PAFs were 43.4% (37.4%-49.0%) by the weighted multiplicative method, and 60.9% (55.7%-65.5%) by the additive method.

### Variation in risk by income

The lowest income group exhibited the highest point prevalence for all risk factors except excessive alcohol consumption and hypertension (Figure 1; Table 4). There was a consistent gradient of decreasing prevalence and both PAFs (both crude and weighted) across increasing income categories for 9 of the 12 risk factors: less education, hearing loss, obesity, smoking, depression, social isolation, physical inactivity, diabetes, and air pollution (Figure 1). Using the weighted multiplicative combined PAF, we estimate that the 12 risk factors accounted for 58.7% of dementia cases in the lowest income group but only 31.8% in the highest income group. The combined additive PAF was 71.3% in the lowest income group and 51.2% in the highest group (Table 4).

**Figure 1.**
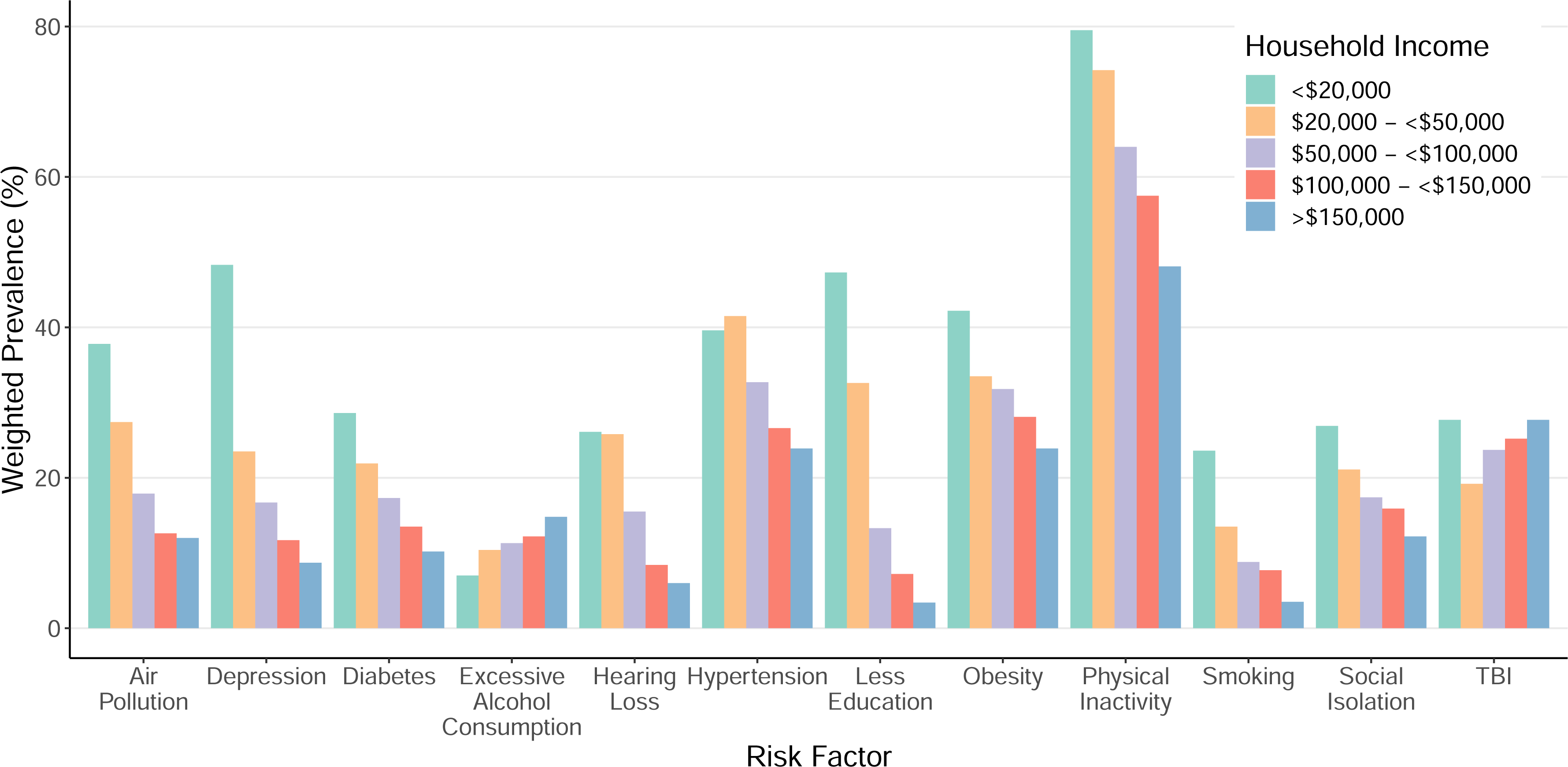
Weighted prevalence of dementia risk factors by household income in CLSA comprehensive cohort at baseline.

**Table 4.**
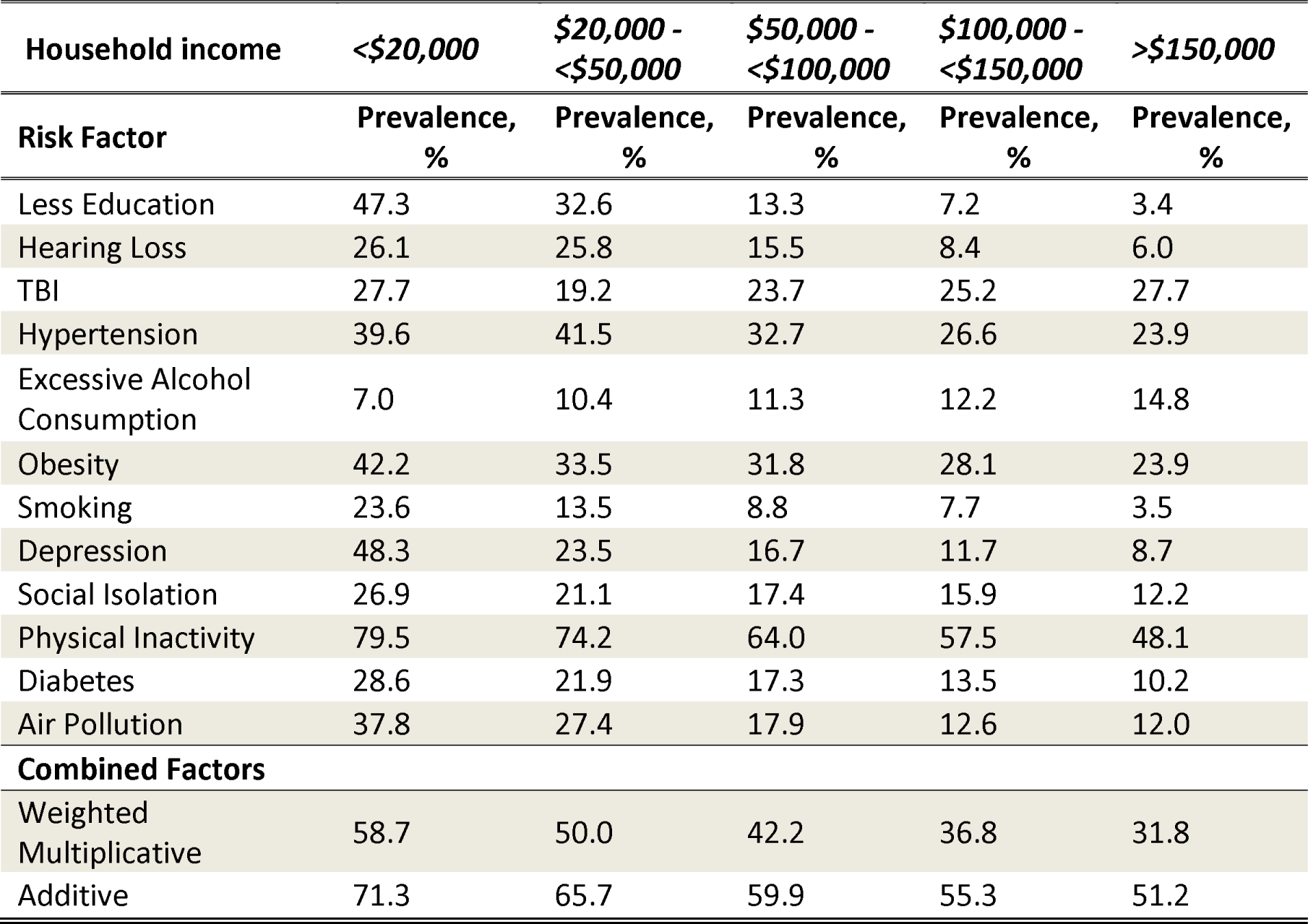
The weighted prevalence and combined population attributable fractions (PAFs) for dementia risk factors by income group in CLSA the comprehensive cohort at baseline.

| Household income | <\$20,000 | \$20,000 - <\$50,000 | \$50,000 - <\$100,000 | \$100,000 - <\$150,000 | >\$150,000 |
| --- | --- | --- | --- | --- | --- |
| Risk Factor | Prevalence, % | Prevalence, % | Prevalence, % | Prevalence, % | Prevalence, % |
| Less Education | 47.3 | 32.6 | 13.3 | 7.2 | 3.4 |
| Hearing Loss | 26.1 | 25.8 | 15.5 | 8.4 | 6.0 |
| TBI | 27.7 | 19.2 | 23.7 | 25.2 | 27.7 |
| Hypertension | 39.6 | 41.5 | 32.7 | 26.6 | 23.9 |
| Excessive Alcohol Consumption | 7.0 | 10.4 | 11.3 | 12.2 | 14.8 |
| Obesity | 42.2 | 33.5 | 31.8 | 28.1 | 23.9 |
| Smoking | 23.6 | 13.5 | 8.8 | 7.7 | 3.5 |
| Depression | 48.3 | 23.5 | 16.7 | 11.7 | 8.7 |
| Social Isolation | 26.9 | 21.1 | 17.4 | 15.9 | 12.2 |
| Physical Inactivity | 79.5 | 74.2 | 64.0 | 57.5 | 48.1 |
| Diabetes | 28.6 | 21.9 | 17.3 | 13.5 | 10.2 |
| Air Pollution | 37.8 | 27.4 | 17.9 | 12.6 | 12.0 |
| **Combined Factors** |  |  |  |  |  |
| Weighted Multiplicative | 58.7 | 50.0 | 42.2 | 36.8 | 31.8 |
| Additive | 71.3 | 65.7 | 59.9 | 55.3 | 51.2 |

### Sensitivity Analysis

The risk factor prevalences and PAFs were broadly similar at follow-up 1 compared to baseline (Table 5). Air pollution dropped from 19.0% at baseline to 13.1% at follow-up 1. Physical inactivity was still the most prevalent risk factor with the highest crude and weighted PAFs. As with the baseline analysis, hypertension and TBI had the next highest crude PAFs, followed closely by obesity. The order of the weighted PAFs changed slighted in follow-up 1 compared to baseline as obesity had a higher weighted PAF than hypertension. The combined PAFs were similar between baseline and follow-up 1.

**Table 5.** The weighted prevalence, relative risk, communality, and population attributable fractions (PAF) of dementia risk factors in the CLSA comprehensive cohort at follow-up 1 (2015-2018, n=27,768).

| **Risk Factor** | **Prevalence, % (95% CI)** | **Relative risk, (95% CI)** | **Communality %** | **Crude PAF, %** | **Weighted PAF, %** |
| --- | --- | --- | --- | --- | --- |
| Less Education | 16.0 (14.7, 17.3) | 1.59 (1.26 – 2.01) | 60 | 8.6 (4.0, 13.9) | 3.4 (1.6, 5.6) |
| Hearing Loss | 16.2 (15.4, 17.1) | 1.94 (1.38 – 2.73) | 72 | 13.2 (5.7, 21.7) | 3.7 (1.6, 6.1) |
| TBI | 24.4 (23.4, 25.5) | 1.84 (1.54 – 2.20) | 80 | 17.0 (11.7, 22.6) | 3.4 (2.3, 4.5) |
| Hypertension | 34.7 (33.7, 35.8) | 1.61 (1.16 – 2.24) | 66 | 17.5 (5.0, 30.0) | 5.9 (1.7, 10.2) |
| Excessive Alcohol Consumption | 11.4 (10.8, 12.1) | 1.18 (1.06 – 1.31) | 75 | 2.0 (0.7, 3.5) | 0.5 (0.2, 0.9) |
| Obesity | 31.3 (30.2, 32.5) | 1.60 (1.34 – 1.92) | 57 | 15.8 (9.5, 22.3) | 6.8 (4.1, 9.6) |
| Smoking | 7.7 (7.1, 8.5) | 1.59 (1.15 – 2.20) | 61 | 4.4 (1.0, 8.5) | 1.7 (0.4, 3.3) |
| Depression | 16.0 (15.1, 17.0) | 1.90 (1.55 – 2.33) | 45 | 12.6 (8.0, 17.5) | 6.9 (4.4, 9.6) |
| Social Isolation | 17.7 (16.7, 18.7) | 1.57 (1.32 – 1.85) | 57 | 9.2 (5.5, 13.2) | 3.9 (2.4, 5.7) |
| Physical Inactivity | 63.7 (62.6, 64.8) | 1.39 (1.16 – 1.67) | 57 | 19.8 (9.1, 30.0) | 8.5 (3.9, 12.9) |
| Diabetes | 19.4 (18.6, 20.4) | 1.54 (1.33 – 1.79) | 59 | 9.5 (6.0, 13.3) | 3.9 (2.4, 5.5) |
| Air Pollution | 13.1 (12.1, 14.1) | 1.09 (1.07 – 1.11) | 31 | 1.2 (0.9, 1.4) | 0.8 (0.6, 1.0) |
| **Combined Factors** | **Combined PAF** |  |  |  |  |
| Weighted Multiplicative | 40.1 (34.5, 45.2) |  |  |  |  |
| Additive | 60.8 (55.5, 65.4) |  |  |  |  |

Avoiding the use of sampling weights noticeably lowered the prevalence estimates of low education, smoking, and air pollution (Table S3). However, the top three risk factors by prevalence and PAF remained the same regardless of weighting, and the multiplicative weighted combined PAF (40.5%) and additive combined PAF (59.2%) were also similar. Analysis stratified by sex also yielded largely similar results (Table S4). Males had a higher prevalence of TBI (29.4% vs. 18.8%) and social isolation (20.2% vs. 15.1%) while females had a higher prevalence of depression (21.7% vs 15.1%).

## Discussion

We examined the prevalence and population attributable fractions of 12 potentially modifiable risk factors for dementia in the Canadian middle-aged and older adult population using data from the Canadian Longitudinal Study on Aging. We found that the prevalence of risk factors varied between 9.6% and 63.8%, with physical inactivity the most prevalent risk factor and smoking the least prevalent. Depending on assumptions regarding the interaction of risk factors, we estimated that between 43.4% and 60.9% of dementia cases among Canadian middle-aged and older adults could be attributed to these 12 risk factors. Nine of the 12 risk factors demonstrated clear gradients across income categories, with higher prevalences of risk factors at lower incomes.

Our work employs a similar methodology to other global and regional studies^6–8;^ however, a novel aspect of our work is that we were able to perform all calculations, except relative risk estimation, within a single national sample. We also used a bootstrapping approach to calculate confidence intervals for PAFs that account for variation in both the prevalence of risk factors and in the relative risks, an improvement over other studies that accounted only for variation within the relative risks^7^ or did not calculate confidence intervals^6^. Despite using a similar methodology, the prevalence and PAFs produced in this study exhibit key differences from those in 2020 Lancet Commission report. While we found the risk factors responsible for the most dementia cases in Canada to be physical inactivity (wPAF=11.6%), depression (7.7%), and hypertension (6.0%), the Lancet Commission identified hearing loss (8.2%), less education (7.1%), and smoking (5.2%) as the most impactful risk factors. Differences in prevalence estimates, a result of different target populations and possibly in the definition of risk factors, are largely responsible for the differences in the weighted PAFs. For example, physical inactivity had just a 11.0% prevalence in the Lancet Commission report, compared to 63.8% in our study and 62.8% in a similar study of adults in the United States ^7^. Other Canadian research supports the high prevalence of physical inactivity we measured in this study^29^. The notable differences in individual PAFs between our study and other international and regional studies are meaningful as they indicate that there are important regional differences in risk factor distribution that can guide the composition and focus of national dementia strategies.

Despite the difference in individual PAFs, the combined PAFs we estimated were similar to those from studies conducted in other jurisdictions. The weighted multiplicative PAF for the 12 risk factors from our study (43%) was highly similar to that of Livingston^22^ (40%) and Lee^7^ (41%). Additionally, our additive combined PAF (61%) is similar to a global estimate (56%)^26^. The differences between the magnitude of the weighted multiplicative and additive combined PAFs are meaningful and ultimately rooted in strong assumptions of the form of interactivity of risk factors within individuals. Interactions between multiple risk factors for dementia is highly complex and the form of the interactions likely vary by each distinct set of risk factors present^26,30,31^. While we believe these combined PAF estimates are useful benchmarks, we have limited evidence on how multiple modifiable risk factors for dementia interact with each other and more research on dementia risk factor interaction could improve future estimation of their combined impact.

The results of this study can be used to inform dementia prevention strategies in Canada. Owning to its high prevalence, physical inactivity had the largest PAF, both crude and weighted. Depression had the second highest weighted PAF, a result of its high relative risk and relatively lower correlation with other risk factors. Depression and dementia have a complex and likely bidirectional relationship, but consensus is that depression is an independent risk factor for dementia that could be targeted for intervention^32,33^. Hypertension had the third highest weighted PAF, although this was meaningfully different from obesity, which had a similar prevalence, relative risk, and communality as hypertension. TBI notably had one of the highest crude PAFs, but relatively lower weighted PAF due to a higher communality value. While our study highlights the risk factors with the greatest impact overall, effective prevention requires a combination of individually tailored and population-level interventions. Evidence supports the effectiveness of individualized, multi-domain interventions to reduce risk factors for dementia^34^. Yet many of the most impactful risk factors have structural etiologies which call for a population-level approach to address^35^.

Notably, we demonstrated gradients of increasing risk with decreasing income across 9 of the 12 modifiable risk factors. This is concordant with observations of a higher incidence of dementia among lower income households and higher combined PAFs of risk factors in lower income countries^36,37^. The etiology of the relationship between socio-economic status and dementia risk is likely multifaceted and related to multiple behavioural, social, cultural, occupational, and environmental mechanisms. The consistently higher prevalence of dementia risk factors in lower income groups suggests that social-economic disparities in dementia incidence are unlikely to be resolved by a prevention focus on single factors. Policies to address the socio-economic disparities directly may be more effective. Future work should further investigate the relationship between socio-economic status and dementia, particularly the degree addressing material deprivation may reduce the prevalence of dementia risk factors and incidence of dementia.

### Limitations

Our study is primarily based on self-reported data, which while common in this field of research may introduce recall and social desirability biases. Compared to the target population, the Comprehensive CLSA cohort was whiter, healthier, and wealthier^11^. However, our use of sampling weights calibrated by education partially addresses this limitation. Individuals with cognitive impairment significant enough to prevent them from giving informed consent were excluded from the CLSA at baseline, and this may have resulted in underestimation of the prevalence of some risk factors. With respect to the PAF calculations, the relative risks used were calculated from an international systematic review and are not specific to Canada. While we strove to algin our risk factor definitions with previous work, the Lancet Commission report from which the relative risk were taken used data from multiple sources and did not always have a single, specific definition.

### Conclusion

We examined the prevalence and population attributable fractions of 12 potentially modifiable risk factors for dementia and estimated that they accounted for between 43.3% and 60.9% of dementia cases among middle-aged and older adults in Canada. Accounting for overlap between risk factors, the factors with the highest overall impact on dementia cases were physical inactivity, depression, and hypertension. The differences in the impact of individual risk factors observed between this study and other international and regional studies highlight the importance of tailoring national and regional dementia strategies to the local distribution of risk factors.

## Data Availability

Data are available from the Canadian Longitudinal Study on Aging (www.clsa-elcv.ca) for
researchers who meet the criteria for access to de-identified CLSA data

## Acknowledgements

This research was made possible using the data/biospecimens collected by the Canadian Longitudinal Study on Aging (CLSA). Funding for the CLSA is provided by the Government of Canada through the Canadian Institutes of Health Research under grant reference: LSA 94473 and the Canada Foundation for Innovation, as well as the following provinces, Newfoundland, Nova Scotia, Quebec, Ontario, Manitoba, Alberta, and British Columbia. This research has been conducted using the CLSA dataset Baseline Comprehensive Version 7, under Application Number 22SP004. The CLSA is led by Drs Parminder Raina, Christina Wolfson, and Susan Kirkland. The opinions expressed in this manuscript are the author’s own and do not reflect the views of the CLSA. SO2 and PM2.5 metrics indexed to DMTI Spatial Inc. postal codes, were provided by CANUE (Canadian Urban Environmental Health Research Consortium). PR holds the Raymond and Margaret Labarge Chair in Optimal Aging and Knowledge Application for Optimal Aging, is the Director of the McMaster Institute for Research on Aging and the Labarge Centre for Mobility in Aging and holds a Tier 1 Canada Research Chair in Geroscience. LG is supported by the McLaughlin Foundation Professorship in Population and Public Health. TLA holds a Tier 1 Canada Research Chair in Healthy Aging.

