## Supplemental Table for "Prevalence and population attributable fractions of potentially modifiable risk factors for dementia in Canada: a cross-sectional analysis of the Canadian Longitudinal Study on Aging"

**Supplementary Tables**

**Table S1. Comparison of risk factor definitions**

| **Risk Factor** | **CLSA definition** | **Lee et al.** ^1^ | **Livingston et al.**^2,3^ |
| --- | --- | --- | --- |
| Less Education | Less than secondary school graduation | Self-reported <12 years of schooling | Lack of secondary school education |
| Hearing Loss | Pure-tone audiometry: Average loss in better ear > 25 dB measured at 0.5, 1, 2, and 4 kHz | Pure-tone mean >25 dB hearing threshold in better ear measured at 0.5, 1, 2, and 4 kHz | 25 dB loss measured by pure-tone audiometry |
| Traumatic Brain Injury | Lifetime history of head injury resulting in loss of consciousness, loss of memory, or dazedness | Self-report of lifetime head injury resulting in loss of consciousness | TBI of all severities |
| Hypertension | Self-reported medical diagnosis of high blood pressure or hypertension | Self-report of taking antihypertensive medications or measured SBP ≥140 mm Hg or measured DBP ≥90 mm Hg | >= 140 SBP; >=130 SBP; use of antihypertensives |
| Excessive alcohol consumption | Self-reported frequency of drinking more than 15 alcoholic drinks for males and 10 for females per week in the past 12 months | Self-report of drinking more than 14 alcoholic drinks (eg, 12 oz of beer, 5 oz of wine, 1.5 oz of spirits) per wk | >21 units/week |
| Obesity | Measured body mass index (BMI) ≥ 30 kg/m^2^ | Measured BMI ≥30 | Body-mass index ≥30 |
| Smoking | Self-reported current smoking (at least one cigarette every day for the past 30 days) | Self-report of current smoking | Current smoking |
| Depression | CESD-10 score ≥ 10 | PHQ-9 score ≥10 | depressive episodes; psychological distress; depressive symptoms; |
| Physical inactivity | Self-reported physical activity of less than 75 minutes of vigorous activity or less than 150 minutes of combined moderate and vigorous activity per week | Self-report of not doing either 75 min/wk of vigorous activity or 150 min/wk of moderate activity or 150 min/wk of moderate/ vigorous activity | Low moderate-to-vigorous physical activity |
| Diabetes | Self-reported medical diagnosis of diabetes, borderline diabetes, or high blood sugar | Self-report of diagnosis or measured fasting plasma glucose ≥126 mg/dL or measured HbA1C ≥6.5% | Diagnosis of diabetes |
| Social isolation | Self-reported frequency of participation in family/friend activities outside of household, religious activities, physical activities with other, educational/cultural activities, association activities, volunteer/charity work, and other recreational activities all less than once per week | Self-report of contact with family, friends, religious organizations, organized groups, or volunteering less than once per month | less social contact; lifelong single; windowed; less social activity; smaller social network |
| Air pollution | Nitrogen dioxide (NO_2_): Annual average ≥ 17 ppb *or*  Fine particle matter (PM2.5): 3-year annual average ≥ 8.8 μg/m3 or  Sulfur dioxide (SO2): 3-year annual average ≥ 5 ppb | NO2 concentration ≥8.7 ppb at census tract level | NO2, PM2.5, CO |

**Table S2: Technical Formulae**

**Abbreviations**

p = Prevalence of risk factor
RR = Casual relative risk of risk factor for all-cause dementia
PAF = Population attributable fraction

**Levin’s Formula for population attributable fraction**^4^

$$\mathrm{PAF}_{j}=\frac{p_{j}({RR}_{j}-1)}{p_{j}({RR}_{j}-1)+1}$$

**Norton et al. formula for weighted population attributable fraction**^5^

$$\begin{aligned} w_{j}=1-\mathrm{communality}_{j} \end{aligned}$$

NB. Communality calculated via performing principal components analysis on tetrachoric correlation matrix of risk factors and extracting components with eigenvalues >1

$$\mathrm{PAF}_{j}=w_{j}\times\mathrm{PAF}_{j}$$

**Barnes and Yaffe formulation for unweighted multiplicative combined population attributable fraction**^6^

$$\mathrm{PAF}_{combined\_mult}=1-\prod_{j=1}^{k} (1-\mathrm{PAF}_{j})$$

**Norton et al. formula for weighted multiplicative combined population attribution fraction**^5^

$$\mathrm{PAF}_{combined\_wmult}=1-\prod_{j=1}^{k} (1-[w_{j}\times\mathrm{PAF}_{j}])$$

**Welberry et al. formula for additive combined population attribution fraction**^7^

$$\begin{aligned} \mathrm{PAF}_{combined\_add}=\frac{\sum_{j=1}^{k} p_{j}({RR}_{j}-1)}{[\sum_{j=1}^{k} p_{j}({RR}_{j}-1)]+1}. \end{aligned}$$

**Table S3.** The unweighted proportions of risk factors for dementia in in CLSA comprehensive cohort at baseline, with associated crude and weighted PAFs

| **Risk Factor** | **Prevalence, % (95% CI)** | **Relative risk, (95% CI)** | **Communality %** | **Crude PAF,  %** | **Weighted PAF, %** |
| --- | --- | --- | --- | --- | --- |
| Less Education | 5.5 (5.2 – 5.7) | 1.59 (1.26 – 2.01) | 60 | 3.1 | 1.4 |
| Hearing Loss | 18.9 (18.5 – 19.4) | 1.94 (1.38 – 2.73) | 72 | 15.1 | 3.9 |
| TBI | 24.2 (23.7 – 24.7) | 1.84 (1.54 – 2.20) | 80 | 16.9 | 4.9 |
| Hypertension | 37.1 (36.6 – 37.7) | 1.61 (1.16 – 2.24) | 66 | 18.5 | 6.6 |
| Excessive Alcohol Consumption | 11.8 (11.5 – 12.2) | 1.18 (1.06 – 1.31) | 75 | 2.1 | 0.4 |
| Obesity | 29.4 (28.8 – 29.9) | 1.60 (1.34 – 1.92) | 57 | 15 | 5.4 |
| Smoking | 6.9 (6.7 – 7.2) | 1.59 (1.15 – 2.20) | 61 | 3.9 | 1.5 |
| Depression | 15.9 (15.5 – 16.3) | 1.90 (1.55 – 2.33) | 45 | 12.5 | 6.8 |
| Social Isolation | 14.1 (13.7 – 14.5) | 1.57 (1.32 – 1.85) | 57 | 7.5 | 3.9 |
| Physical Inactivity | 63.5 (62.9 – 64.0) | 1.39 (1.16 – 1.67) | 57 | 19.8 | 11.5 |
| Diabetes | 17.7 (17.3 – 18.3) | 1.54 (1.33 – 1.79) | 59 | 8.7 | 3.6 |
| Air Pollution | 7.9 (7.6 – 8.2) | 1.09 (1.07 – 1.11) | 31 | 0.7 | 0.3 |
| **Combined Factors** | **Combined PAF** |  |  |  |  |
| Weighted Multiplicative | 40.5 |  |  |  |  |
| Additive | 59.2 |  |  |  |  |

| **Table S4.** The weighted prevalence and PAFs of dementia risk factors by sex in the CLSA comprehensive cohort at baseline. | | | | | | | |
| --- | --- | --- | --- | --- | --- | --- | --- |
|  | ***Male*** | | | | ***Female*** | | |
| **Risk Factor** | | **Prevalence %** | **PAF, %** | **wPAF %** | **Prevalence %** | **PAF, %** | **wPAF %** |
| Less Education | | 16.9 | 9.1 | 4.2 | 17.8 | 9.5 | 4.4 |
| Hearing Loss | | 17.4 | 14.1 | 3.7 | 14.1 | 11.7 | 3.0 |
| TBI | | 29.4 | 19.8 | 5.7 | 18.8 | 13.7 | 3.9 |
| Hypertension | | 34.8 | 17.5 | 6.3 | 30.9 | 15.9 | 5.7 |
| Excessive Alcohol Consumption | | 12.7 | 2.2 | 0.4 | 10.2 | 1.8 | 0.3 |
| Obesity | | 31.2 | 15.8 | 5.7 | 30.4 | 15.4 | 5.6 |
| Smoking | | 9.5 | 5.3 | 2.1 | 9.6 | 5.4 | 2.1 |
| Depression | | 15.1 | 11.9 | 6.4 | 21.7 | 16.4 | 8.8 |
| Social Isolation | | 20.2 | 10.3 | 5.4 | 15.1 | 7.9 | 4.1 |
| Physical Inactivity | | 60.8 | 19.2 | 11.1 | 66.5 | 20.6 | 11.9 |
| Diabetes | | 19.3 | 9.5 | 3.9 | 15.6 | 7.8 | 3.2 |
| Air Pollution | | 18.8 | 1.7 | 0.8 | 19.3 | 1.7 | 0.8 |
| **Combined Factors** | |  |  |  |  |  |  |
| Weighted Multiplicative | | 43.7 |  |  | 42.8 |  |  |
| Additive | | 61.5 |  |  | 59.7 |  |  |
